# Identification of patients at risk for pancreatic cancer in a 3-year timeframe based on machine learning algorithms

**DOI:** 10.1101/2023.08.03.23293555

**Authors:** Weicheng Zhu, Yindalon Aphinyanaphongs, Fay Kastrinos, Diane M. Simeone, Mark Pochapin, Narges Razavian, Tamas Gonda

## Abstract

**Importance:** Early detection of pancreatic cancer (PC) remains challenging largely due to the low population incidence and few known risk factors. However, screening in at-risk populations and detection of early cancer has the potential to significantly alter survival.

**Objective:** In this study, we aim to develop a predictive model to identify patients at risk for developing new-onset PC at two and a half to three year time frame

**Data Sources:** We used the Electronic Health Records (EHR) of a large medical system from 2000 to 2021 (N=537,410). The EHR data analyzed in this work consists of patients’ demographic information, diagnosis records, and lab values, which are used to identify patients who were diagnosed with pancreatic cancer and the risk factors used in the machine learning algorithm for prediction.

**Results:** We identified 73 risk factors of pancreatic cancer with the Phenome-wide Association Study (PheWAS) on a matched case-control cohort. Based on them, we built a large-scale machine learning algorithm based on EHR. A temporally stratified validation based on patients not included in any stage of the training of the model was performed. This model showed an AUROC at 0.742 [0.727, 0.757] which was similar in both the general population and in a subset of the population who has had prior cross-sectional imaging. The prevalence of pancreatic cancer in those in the top 5 percentile of the risk score was 6 folds higher than the general population.

**Conclusion:** Our model leverages data extracted from a 6-month window of time in the electronic health record to identify patients at nearly 6-fold higher than baseline risk of developing pancreatic cancer 2.5 to 3 years from evaluation. This approach offers an opportunity to define an enriched population entirely based on static data, where current screening may be recommended.

## 1. Introduction

Despite the relatively low incidence of pancreatic cancer of 13.2 per 100,000, it now ranks as the third leading cause of cancer death [1]. While advances continue to be made in therapy most patients are diagnosed with advanced and incurable disease. Early detection offers a critical path toward improved overall survival. Pancreatic cancer diagnosed at an early stage has been associated with significantly greater survival and when PC is detected in a screening program, it is associated with significantly better outcomes [2]. However, due to the low prevalence and current lack of effective non-invasive screening modalities, population-level screening has not been endorsed by the US Preventative Services Task Force. Currently, most patients who are enrolled in a screening program are identified based on family history or known germline mutation status (referred to as high-risk individuals; HRI [3]) or after an incidental diagnosis of a cystic neoplasm. However, only 10-20% of patients diagnosed with PC would have been eligible for screening based on family history or germline mutations and no more than 15% of cancers arise from cystic lesions. Therefore, the majority of patients ultimately diagnosed with PC would not have been offered screening according to current practices. In addition, significant disparities exist in the recognition and testing of HRI, which further decreases the efficacy of current screening recommendations [4].

There are several risk factors that are associated with pancreatic cancer, but none have sufficiently high odds to help identify a population at risk. Risk associations were developed based on the combination of several of these risk factors and their dynamic change during a period preceding the diagnosis of cancer [23, 24]. In addition, some of these risk factors were specifically applied to cohorts with existing pancreatic conditions that are associated with PDAC such as chronic pancreatitis or cystic neoplasms [25, 26, 27]. Recent studies have highlighted the opportunities offered my machine learning approaches to mining of the EHR data and the unselected general population. A recent transformer model based study, trained on a Danish cohort and validated in the US Veterans Affair Health System [28], used a prediction time of 0-5 years and another US-based study built a prediction model with a more defined but relatively short time frame between evaluation and diagnosis (180-365 days) [29]. Another recent study evaluated an even shorter lead time of 3 months[30]. In comparison, we chose a longer 2.5-3 year timeframe between prediction time and pancreatic cancer diagnosis based on biologic and epidemiologic data as well as greatest potential for clinical impact on survival. Our goal was to primarily evaluate a longer and fixed time frame that we believed was biologically reasonable and clinically impactful. We chose 2.5-3 years between the prediction time and pancreatic cancer diagnosis. In addition, instead of a black box approach we first perform a statistically rigorous PheWAS feature selection step on a carefully matched PC case-control sub-cohort, and then train the model based on the selected features on the full dataset. Furthermore, we validate our model on a heldout cohort that is also temporally different compared to the training and development cohort. This further strengthens the generalizability estimate of the model, given that the practice of medicine and documentation changes, and major world events such as COVID-19 pandemic create a large distribution shift in the data distribution over time.

We used the Electronic Health Record (EHR) of our large medical system to identify patients who were diagnosed with pancreatic cancer and had a presence for three or more years in the health record preceding the event. We identified a matched control cohort (N=7,728) based on comorbidities and length of presence of database as our pancreatic cancer cohort (N=1,923), using the k-nearest neighbor algorithm. We then used phenome-wide association methodology [5,6] to identify diagnosis codes (phenotypic traits) that are associated with the target outcome (pancreatic cancer). 73 diagnosis codes and 5 lab values were found significantly associated with PC onset. Finally, a risk model was trained and evaluated based on the identified risk factors on the overall patient population eligible for predictive modeling (N=996,384). We developed our model on a cohort of individuals present in the database up to 2015 (N=527,027) and validated the model of a (new patient, not overlapping with the development set) cohort between 2015 to 2022 (N=469,357). This temporally-stratified validation mimics real-world scenarios of training and deploying the model.

Our objective was to develop and validate a model that can identify at risk individuals from the general population. In over 70 variables, our variables include known and previously unrecognized risk factors for pancreatic cancer, and our model identifies a population where the baseline prevalence is nearly six-fold higher than the general population. We believe this risk stratification can help identify a sufficiently enriched patient population where current screening approaches can be recommended and this static EHR-based approach can offer an opportunity to readily identify these individuals.

## 2. Methods

### 2.1 Study Design

This predictive modeling study was performed based on electronic health records (EHR) at New York University (NYU) Langone Health. This study followed the Strengthening the Reporting of Observational Studies in Epidemiology (STROBE) reporting guideline for cohort studies. The overview of our approach is described in Figure 1. We first identify patients with pancreatic cancer and at least three years of records prior to their pancreatic cancer disease onset (N=1,923). We then matched these patients to a set of control patients (N=7,728) based on demographic features and based on presence in the database for at least three years. We used phenome-wide association methodology on this matched case-control set, to identify factors that are statistically significantly associated with pancreatic cancer (73 diagnosis codes and 5 lab values were selected). Our training or discovery dataset was based on a patient population who last visited NYU before or in 2015 (N=527,027). Our performance results are then reported on a temporally distinct validation cohort, of a new set of patients who visited NYU after 2015 (N=469,357).

**Figure 1.**
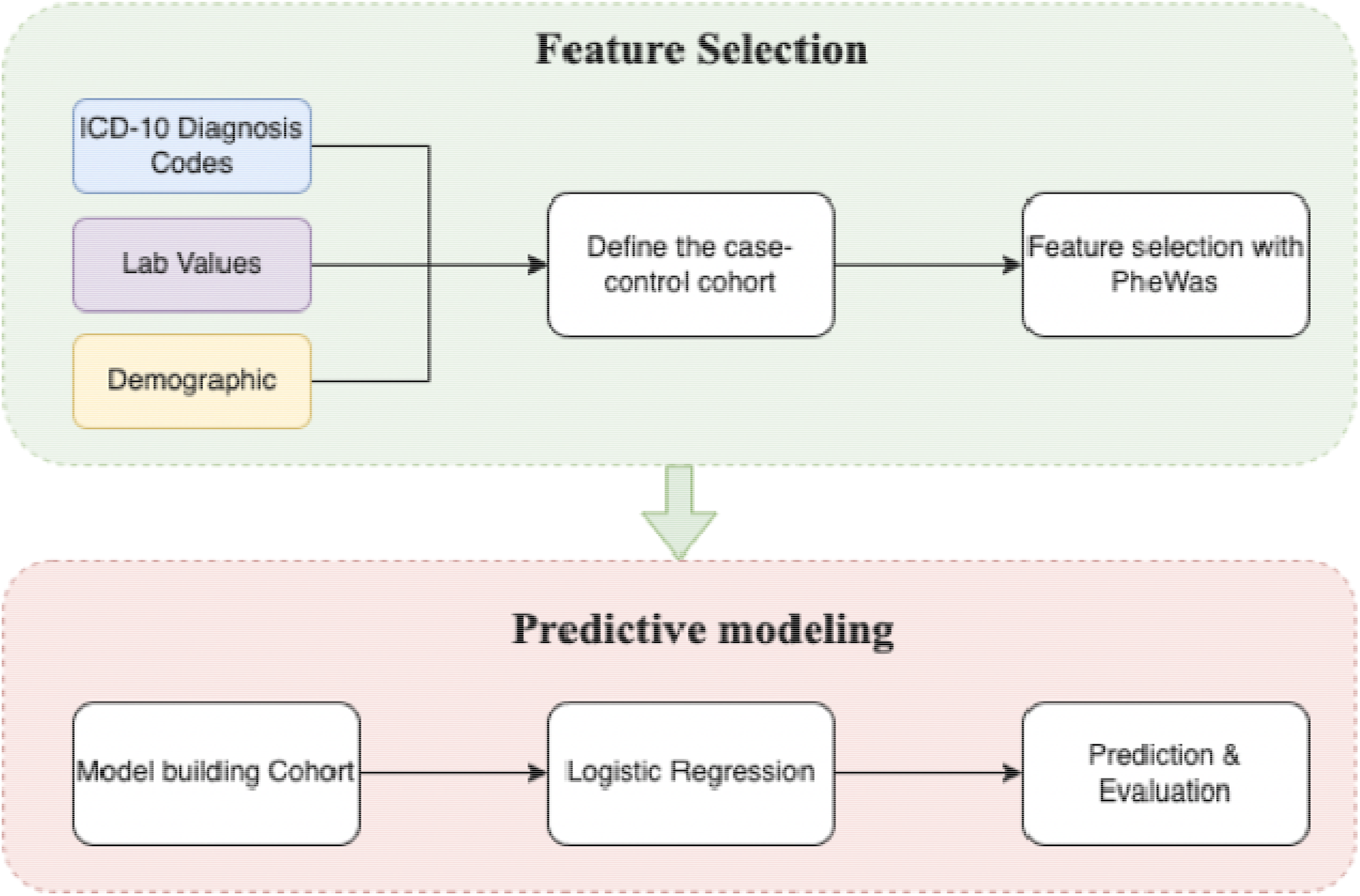
The pipeline of our model.

### 2.2 Overall Population, Pancreatic Cancer Definition, and Definition of Included Variables

We identified individuals with at least 3 years of continuous presence in the EHR between January 2000 and January 2021 in the NYU Langone Health system. The EHR data analyzed in this work consists of patients’ demographic information, diagnosis records, and lab values. Diagnosis of pancreatic adenocarcinoma was identified by the diagnostic codes using encounter and billing diagnoses according to the criteria in Supplementary Table A1.

### 2.3 Feature Selection with Phenome-wide Association (PheWAS) Analysis

In this section, we introduce feature selection with PheWAS. We first defined the inclusion criteria of positive and negative samples for pancreatic cancer. From them, we matched a case-control cohort under the control of other confounding factors. Then a PheWAS analysis was conducted on the case cohort against the control cohort to select features.

#### Inclusion Criteria and Prediction Time for Pancreatic Cancer Patients

The timeline of developing pancreatic cancer (PC) is critical in this study. The date of PC diagnosis was defined as the first date at which the PC criteria was observed in the patient records. To predict PC within a 3-year timeframe, we only included PC patients that had at least 3 years’ worth of data prior to the first PC diagnosis time. We then defined a date for each patient, called *<u>prediction time</u>*, assuming that we made predictions on that date based on historical data prior to this date. Patients without any records prior to the prediction time were excluded.

#### Inclusion Criteria and Prediction Time for Pancreatic Cancer-free Patients

For each patient without Pancreatic Cancer, a random encounter date was selected as the “prediction time”. Inclusion criteria were defined as having 3 years follow-up at NYU Langone after that “prediction time”. Patients without any history prior to the prediction time were excluded.

#### Included Variables for PheWAS Analysis

Disease records from a 6-month window prior to the “prediction time”, in addition to the *latest* lab and physiological data (unrestricted by time, any time prior to the “prediction time”) were used to construct variables for the PheWAS stage. Previous diseases of patients prior to

“prediction time” were represented as 19,304 ICD-10 codes derived from the EHR. The following 10 lab results or physiological data of patients were included, based on the clinical potential to be a risk factor of PC [32, 33, 34]. These lab values include A1C, Glucose, Hemoglobin, Albumin, Lipase, Amylase, AST, CA19-9, and BMI. We binarized lab values referring to the standard normal ranges (Supplementary Table A2), and bucketed BMI into underweight (BMI<18.5), normal (BMI 18.5 −25), overweight (BMI 25 - 30), and obese (BMI >30).

#### Phenome-wide Association Analysis

We introduce Phenome-wide association study (PheWAS) [5,6] to conduct feature selection among thousands of diagnosis variables and lab values. PheWAS is designed to measure the association between a set of phenotypes and target outcomes, with the aim of identifying links between known phenotypic traits (including molecular, biochemical, cellular, and clinical diagnoses) and the target outcomes of interest. However, PheWAS can be influenced by various confounding variables, such as age and sex. To address this, we conducted a case-control subcohort analysis to match each pancreatic cancer (PC) patient with a non-PC patient based on age, sex, length of history in the EHR, and the number of ICD codes. We matched each included PC patient with a non-PC patient using the k-nearest neighbor algorithm, and we used a 1:4 case-to-control ratio. We used the PheWAS approach to test the association between the new onset PC and the included 19,304 diagnosis variables and 10 lab values and physiological measures. Statistically significant variables were used in the subsequent predictive modeling.

#### Statistical Analysis

In the PheWAS analysis, we reported the odds ratios (unadjusted) and the corresponding p-values of the association between the condition and new onset PC in a three-year follow-up. For binary disease indicators (documented at ICD-10), the unadjusted odds ratios (UOR) and p-values of the association were computed based on the Chi-square test; for bucketed numerical values, the UORs were computed on the sub-cohort of patients who had undergone the lab test. The p-values and confidence intervals of the UORs were computed using the proportion z-test. Correction for multiple hypothesis testing was performed using Bonferroni correction, with the level of significance (*) adjusted after Bonferroni correction. Statistically significant diagnosis variables were selected based on the p-values with the threshold of 0.01 after correction for multiple hypothesis testing.

### 2.4 Predictive Modeling

#### Inclusion Criteria for Predictive Modeling with Temporal Cohort Stratification

Different inclusion criteria of patients and included variables are introduced to simulate the real-world predictive operation. We used temporal stratification to separate the development and validation set because we can only use the past data for model development in practice. The development cohort for predictive model development included patients older than 40 years (at their last visit), with or without PC whose last record in EHR was in 2015, who matched the following inclusion criteria: For PC patients, we required at least 3 years of data prior to the first date where any record of PC was observed. For PC-free patients, any patient with at least 3 years of records were included. For PC patients, 2.5 years prior to the first PC record was selected as the prediction time. For PC-free patients, “prediction time” was selected as 3 years prior to their last visit. An independent cohort of new patients, not used during the development stage, was similarly constructed, from patients whose last visit happened between 2015 and 2022.

#### Included Variables for Predictive Modeling

During the predictive modeling, variables selected as significant during the PheWAS stage were constructed for all included patients from all records prior to the “prediction time”.

#### Model Development

We built two predictive models, based on regularized logistic regression models with elastic net based regularization [7]. The first model was trained on the full population that is older than 40 years old (N=527,027). The second model was trained and validated only on patients with no known or documented pancreatic conditions or cross-sectional imaging. The prediction models were regularized Logistic Regression and we tuned the regularization penalty by elastic net (the combination of L1 and L2 regularization). During the development stage, we split the development data set into a training set and held-out validation and performed cross-validation. We then reported results on the temporally separate (new patient) held-out cohort described in the previous section.

##### Statistical Analysis

In predictive modeling, we evaluated the model performance on the held-out validation set with Areas Under the Receiver Operating Characteristic Curve (AUROC). Additional metrics, including Positive Predictive Value (PPV), Negative Predictive Value (NPV), sensitivity, and specificity were computed at different probability thresholds on the predictions, to facilitate the identification of the optimal threshold for screening. We also reported the fully adjusted odds ratios (OR) and p-values of the variables associated with PC onset based on the logistic regression analysis.

### 2.5 Sensitivity Analysis

The accuracy of our data relies heavily on the accuracy of the use of diagnosis codes, which is a manual chart review of randomly selected cases and controls we found to be approximately 80% accurate. Therefore, to examine the stability of our model on data noise, we conducted a sensitivity analysis by retraining the model based on simulated data where we randomly flipped the labels of 20% cancer samples and the same number of cancer-free samples. We reported the mean and standard deviation 10 times of such experiments to analyze incorrect EHR labels’ impact.

## 3. Results

### 3.1 Identification of Cohorts for PheWAS and Predictive Modeling

537,410 patients were eligible to be included, where 1,932 patients with PC were matched to 7,728 PC-free patients based on demographics and length of presence in the database. The characteristics of the case and control cohort are summarized in Table 1.

**Table 1.**
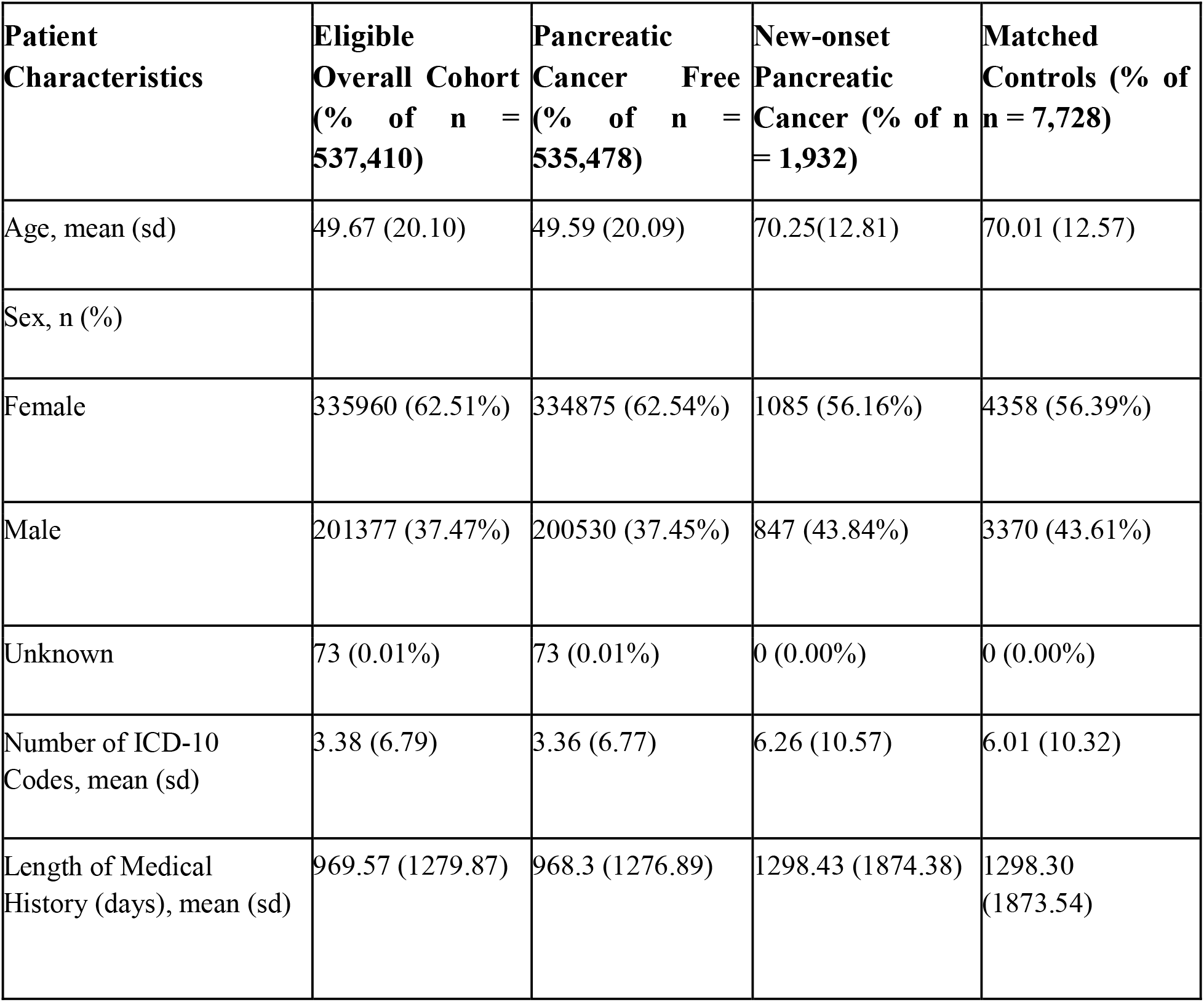
Demographic and overall characteristics of all eligible individuals, pancreatic cancer patients, all cancer-free individuals and matched cohorts, used during the PheWAS analysis.

| **Patient Characteristics** | **Eligible Overall Cohort (% of n = 537,410)** | **Pancreatic Cancer Free (% of n = 535,478)** | **New-onset Pancreatic Cancer (% of n = 1,932)** | **Matched Controls (% of n = 7,728)** |
| --- | --- | --- | --- | --- |
| Age, mean (sd) | 49.67 (20.10) | 49.59 (20.09) | 70.25(12.81) | 70.01 (12.57) |
| Sex, n (%) |  |  |  |  |
| Female | 335960 (62.51%) | 334875 (62.54%) | 1085 (56.16%) | 4358 (56.39%) |
| Male | 201377 (37.47%) | 200530 (37.45%) | 847 (43.84%) | 3370 (43.61%) |
| Unknown | 73 (0.01%) | 73 (0.01%) | 0 (0.00%) | 0 (0.00%) |
| Number of ICD-10 Codes, mean (sd) | 3.38 (6.79) | 3.36 (6.77) | 6.26 (10.57) | 6.01 (10.32) |
| Length of Medical History (days), mean (sd) | 969.57 (1279.87) | 968.3 (1276.89) | 1298.43 (1874.38) | 1298.30 (1873.54) |

### 3.2 PheWAS Analysis Results

After performing PheWAS analysis on the matched case-control, based on 19,304 disease records and 10 laboratory tests or physiological signs (BMI, A1C, Hemoglobin, fasting glucose, CA19-9, Lipase, Amylase, AST, ALT, Albumin), we identified 73 diagnosis codes and 5 lab values that showed significant association with PC onset. Table 2 shows the top 20 variables and Suppl Table A3 shows the comprehensive list. Most of the features have been associated with pancreatic cancer in the past, however, we included all of these features in predictive modeling. We noted that several of the highly significant features could only have been recognized in patients with either a past medical history of pancreatic diseases or cross-sectional imaging (e.g. pancreatic cyst). As this represented a minority but a potentially distinct and higher-risk population, we developed a model for all eligible individuals (Model 1) and a separate model for those without pancreatic disease or cross-sectional imaging in the time period of feature selection (Model 2).

**Table 2.** The top 20 variables that are associated with the development of pancreatic cancer. The unadjusted odds ratios on the case-control cohort and the unadjusted odds ratios of the two models on the overall cohort are shown. *Note that the statistically significant features after Bonferroni correction are marked (*** means corrected p-value < 0.001; ** means corrected p-value < 0.05; * means corrected p-value < 0.1)

| **Diagnosis Features** | **Control (%)** | **Case (%)** | **P-value** | **Unadjusted Odds Ratio**<br>**[95% Confidence Interval]** |
| --- | --- | --- | --- | --- |
| Disease of pancreas, unspecified (ICD10 K86.9) | 2(0.02%) | 11(0.53%) | 5.61E-09 ** | 22.11 [4.90, 99.83] |
| Malignant neoplasm of transverse colon (ICD10 C18.4) | 1(0.01%) | 5(0.24%) | 1.05E-04 | 20.05 [2.34, 171.68] |
| Pseudocyst of pancreas (ICD10 K86.3) | 6(0.07%) | 16(0.77%) | 6.04E-10 ** | 10.74 [4.20, 27.48] |
| Hypertrophy of breast (ICD10 N62) | 4(0.05%) | 9(0.43%) | 8.98E-06 ** | 9.03 [2.78, 29.37] |
| Neoplasm of unspecified behavior of digestive system (ICD10 D49.0) | 8(0.10%) | 16(0.77%) | 1.05E-08 ** | 8.05 [3.44, 18.84] |
| Other specified diseases of pancreas (ICD10 K86.89) | 11(0.13%) | 15(0.72%) | 1.50E-06 ** | 5.49 [2.52, 11.96] |
| Genetic susceptibility to other malignant neoplasm (ICD10 Z15.09) | 10(0.12%) | 13(0.62%) | 1.17E-05 ** | 5.23 [2.29, 11.94] |
| Genetic susceptibility to malignant neoplasm of breast (ICD10 Z15.01) | 9(0.11%) | 11(0.53%) | 8.97E-05 | 4.91 [2.03, 11.86] |
| Malignant neoplasm of unspecified ovary (ICD10 C56.9/57.00) | 6(0.07%) | 7(0.33%) | 2.27E-03 | 4.68 [1.57, 13.94] |
| Acute pancreatitis without necrosis or infection, unspecified (ICD10 K85.90) | 7(0.08%) | 7(0.33%) | 4.98E-03 | 4.01 [1.41, 11.45] |
| Family history of malignant neoplasm, unspecified (ICD10 Z80.9) | 18(0.22%) | 15(0.72%) | 2.51E-04 | 3.35 [1.69, 6.66] |
| Primary osteoarthritis, unspecified hand (ICD10 M19.049) | 10(0.12%) | 8(0.38%) | 9.46E-03 | 3.21 [1.26, 8.14] |
| Malignant neoplasm of unspecified kidney, except renal pelvis (ICD10 C64.9) | 14(0.17%) | 10(0.48%) | 7.89E-03 | 2.87 [1.27, 6.46] |
| Cerebral infarction due to unspecified occlusion or stenosis of unspecified cerebral artery (ICD10 I63.50) | 24(0.29%) | 15(0.72%) | 3.88E-03 | 2.51 [1.31, 4.79] |
| Malignant neoplasm of unspecified site of unspecified female breast (ICD10 C50.919) | 101(1.21%) | 43(2.06%) | 2.89E-03 | 1.72 [1.20, 2.46] |

| **Lab Value Features** | **Control (%)** | **Case (%)** | **P-value** | **Unadjusted Odds Ratio**<br>**[95% Confidence Interval]** |
| --- | --- | --- | --- | --- |
| Lipase > 234.0 | 29(12.61%) | 20(25.64%) | 6.54E-03 | 2.39 [1.26 , 4.53] |
| Glucose > 126.0 | 208(29.80%) | 106(46.70%) | 3.01E-06 ** | 2.06 [1.52 , 2.81] |
| Obesity (BMI > 30) | 1136(28.32%) | 291(38.70%) | 1.17E-08** | 1.60 [1.36 , 1.88] |
| Type II Diabetes<br>(E11.9 + A1C) | 459(5.49%) | 164(7.84%) | 4.71E-05 * | 1.47 [1.22, 1.76] |
| overweight (BMI 24~30) | 1294(32.25%) | 280(37.23%) | 7.70E-03 | 1.25 [1.06 , 1.47] |

### 3.3 Performance of the Prediction Model

For the first model, we trained and evaluated the regularized logistic regression in Model 1 between 2000 and 2021 (N=996,384). As described in the Methods section, this cohort was temporally stratified into training and (temporally distinct, new patients) held-out validation set. The characteristics of the training set and the held-out validation set are summarized in Table 3.

**Table 3.**
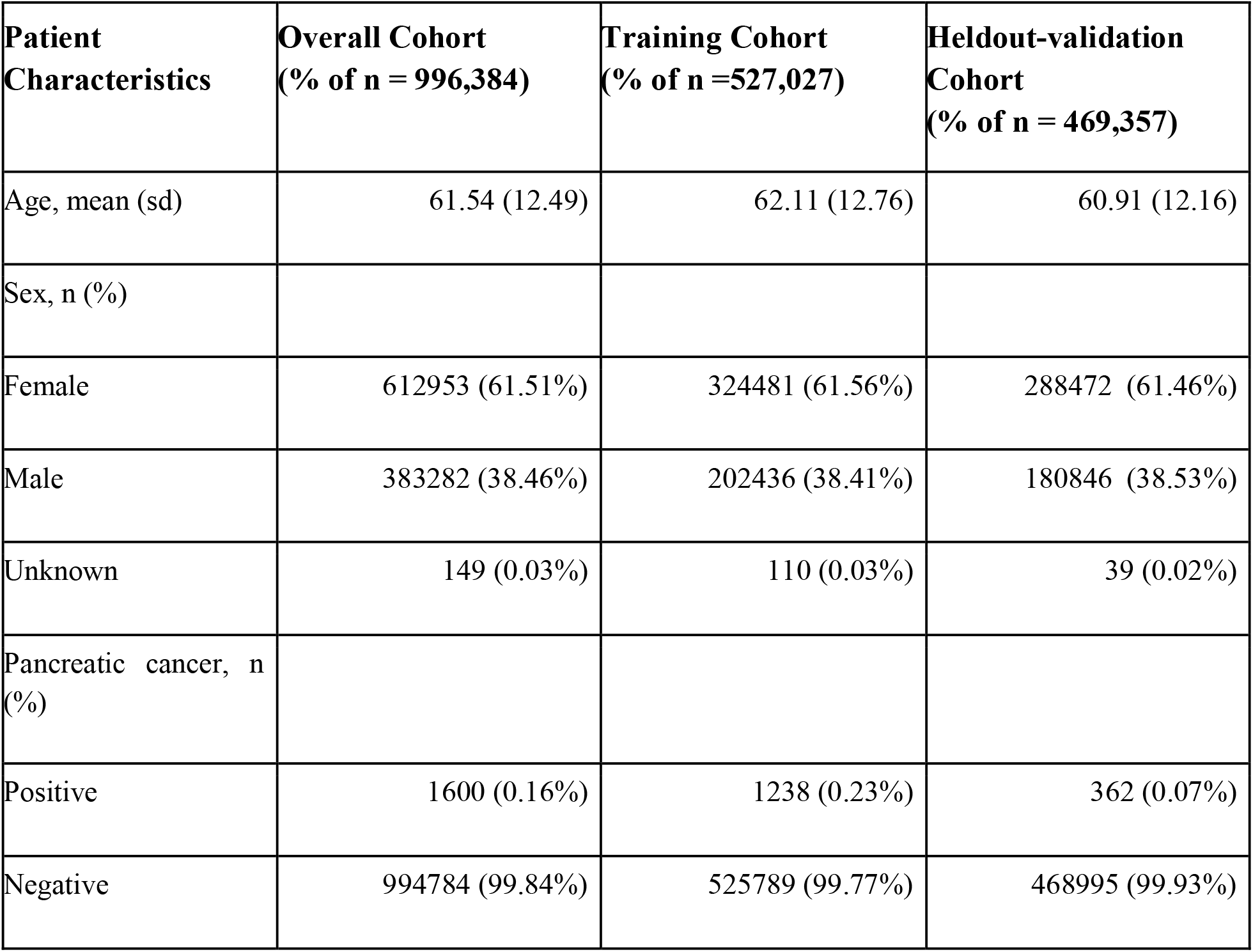
The characteristic table of the overall cohort for predictive modeling, as well as training and test set separately.

**Table 4.** The coefficients and p-values of the features in the logistic regression model.

| **Description** | **P-value** | **Adjusted odds Ratio**<br>**[95% Confidence Interval]** |
| --- | --- | --- |
| Other specified diseases of pancreas (ICD K86.89) | 0.00E+00 *** | 6.429 [4.734, 8.124] |
| Neoplasm of unspecified behavior of digestive system (ICD D49.0) | 6.99E-08 *** | 4.106 [2.243, 5.970] |
| Acute pancreatitis without necrosis or infection, unspecified (ICD K85.90) | 8.44E-14 *** | 4.019 [1.568, 6.470] |
| Malignant neoplasm of unspecified kidney, except renal pelvis (ICD C64.9) | 2.00E-15 *** | 3.947 [2.634, 5.259] |
| Primary osteoarthritis, unspecified hand (ICD M19.049) | 1.39E-10 *** | 2.868 [1.984, 3.752] |
| Family history of malignant neoplasm, unspecified (ICD Z80.9) | 2.22E-16 *** | 2.768 [2.136, 3.399] |
| Genetic susceptibility to other malignant neoplasm (ICD Z15.09) | 0.00E+00 *** | 2.159 [0.597, 3.720] |
| Cyst of pancreas (ICD K86.2) | 2.44E-15 *** | 1.924 [1.127, 2.720] |
| Glucose > 126.0 | 1.45 E-04 * | 1.910 [1.223, 2.596] |
| Malignant neoplasm of unspecified site of unspecified female breast (ICD C50.919) | 0.00E+00 *** | 1.880 [1.660, 2.100] |
| Type 2 diabetes mellitus without complications (ICD E11.9) | 0.00E+00 *** | 1.876 [1.749, 2.002] |
| Personal history of poliomyelitis (ICD Z86.12) | 5.34E-02 | 1.797 [0.996, 2.597] |
| Cerebral infarction due to unspecified occlusion or stenosis of unspecified cerebral artery (ICD I63.50) | 5.50E-03 | 1.780 [1.119, 2.442] |
| Genetic susceptibility to malignant neoplasm of breast (ICD Z15.01) | 1.40E-05 ** | 1.758 [0.714, 2.801] |
| Pseudocyst of pancreas (ICD K86.3) | 4.41E-02 | 1.511 [0.871, 2.151] |
| Malignant neoplasm of unspecified fallopian tube (ICD C57.00) | 4.87E-08 *** | 1.427 [1.241, 1.613] |
| Disease of pancreas, unspecified (ICD K86.9) | 7.97E-04 * | 1.415 [1.135, 1.696] |
| Obesity (BMI > 30) | 9.39E-02 | 1.238 [0.945, 1.531] |
| Lipase > 234.0 | 5.28E-01 | 1.111 [0.790, 1.431] |
| Overweight (BMI 24~30) | 6.10E-01 | 1.087 [0.779, 1.395] |
| A1C > 6.5 | 4.76E-01 | 1.058 [0.905, 1.211] |

**Table 5.** Details on sensitivity, specificity, positive predictive values (NPV), and negative predictive values (NPV) at different risk percentiles based on model 1.

| Risk percentiles | Sensitivity (%) | Specificity (%) | PPV (%) | NPV (%) |
| --- | --- | --- | --- | --- |
| 1% | 6.78 [6.13, 7.43] | 99.01 [99.01, 99.01] | 0.65 [0.58, 0.72] | 99.93 [99.92, 99.93] |
| 5% | 18.36 [16.82, 19.89] | 95.01 [95.01, 95.01] | 0.34 [0.31, 0.37] | 99.92 [99.92, 99.92] |
| 10% | 29.14 [27.70, 30.58] | 90.02 [90.02, 90.02] | 0.28 [0.26, 0.29] | 99.93 [99.92, 99.93] |
| 20% | 42.71 [40.66, 44.76] | 80.02 [80.02, 80.03] | 0.19 [0.19, 0.20] | 99.93 [99.92, 99.94] |

Figure 2 shows the Receiver Operating Characteristic (ROC) curve of Model 1. The Areas Under the ROC curve (AUROC) was 0.742 [0.727, 0.757] and this was not significantly different than the performance in Model 2 (Suppl Figure B1). As the prevalence of pancreatic cancer is low, the Positive Predictive Value (PPV) is crucial for selecting a reasonable threshold for screening. The PPV of the model at different predicted risk levels is shown in Figure 3. Screening the top-1 and top-5 percent of the patients with the highest risks would achieve around 7 and 4 times higher PPV, respectively, than the general patient population in the EHR. We also compared this to the PPV in individuals with Type II Diabetes (T2D; defined as either having any A1c > 6.5, or an E11.9 diagnosis code) based on published risk assessment and found that this model would have 5 times higher PPV than T2D. This implies that the model provides a more efficient tool for identifying high-risk patients.

**Figure 2.**
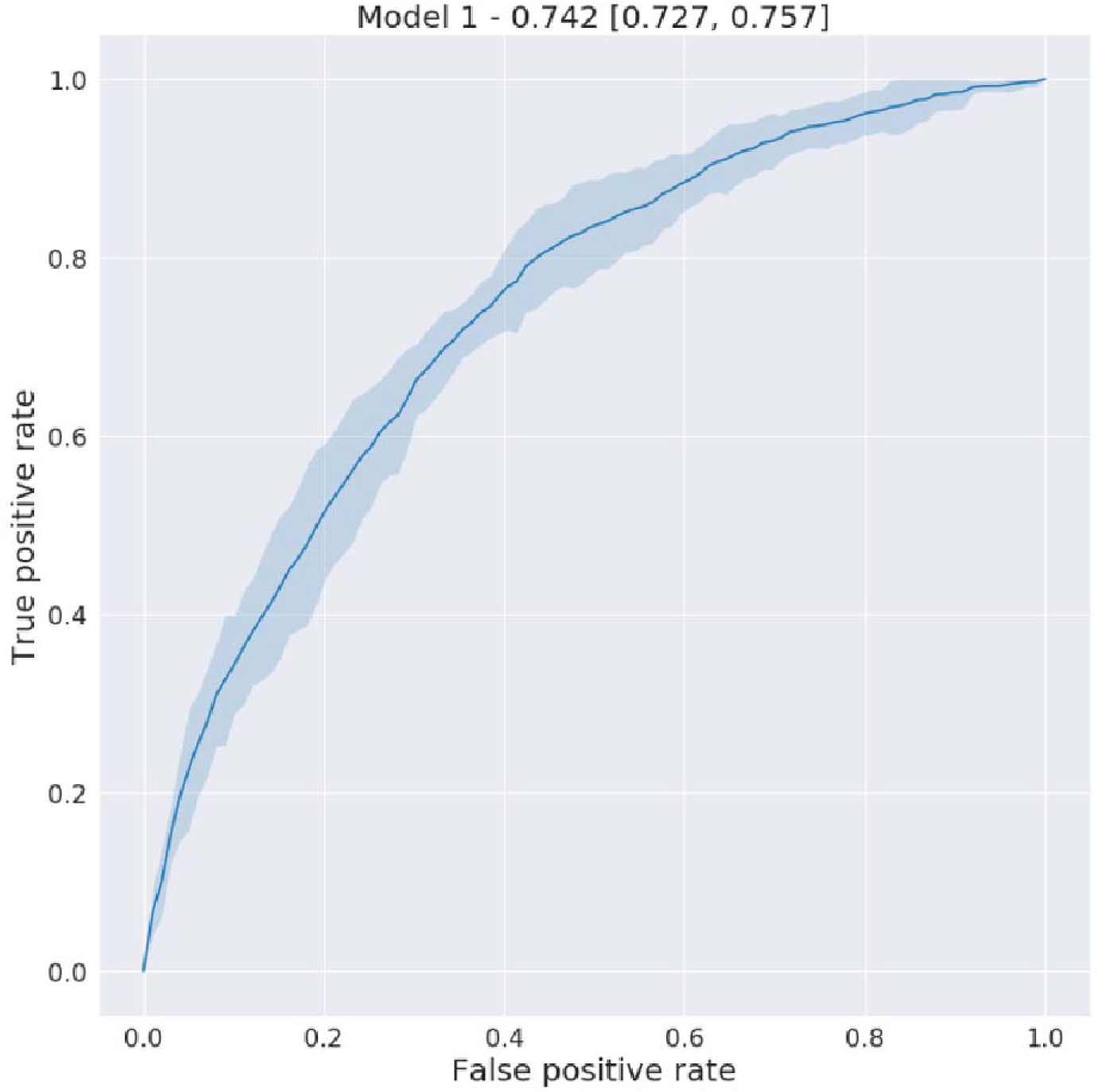
The Receiver Operating Characteristics Curve (AUROC 0.742 [0.727, 0.757]) on the held-out validation set to predict new-onset pancreatic cancer in a 3-year follow-up.

**Fig 3.**
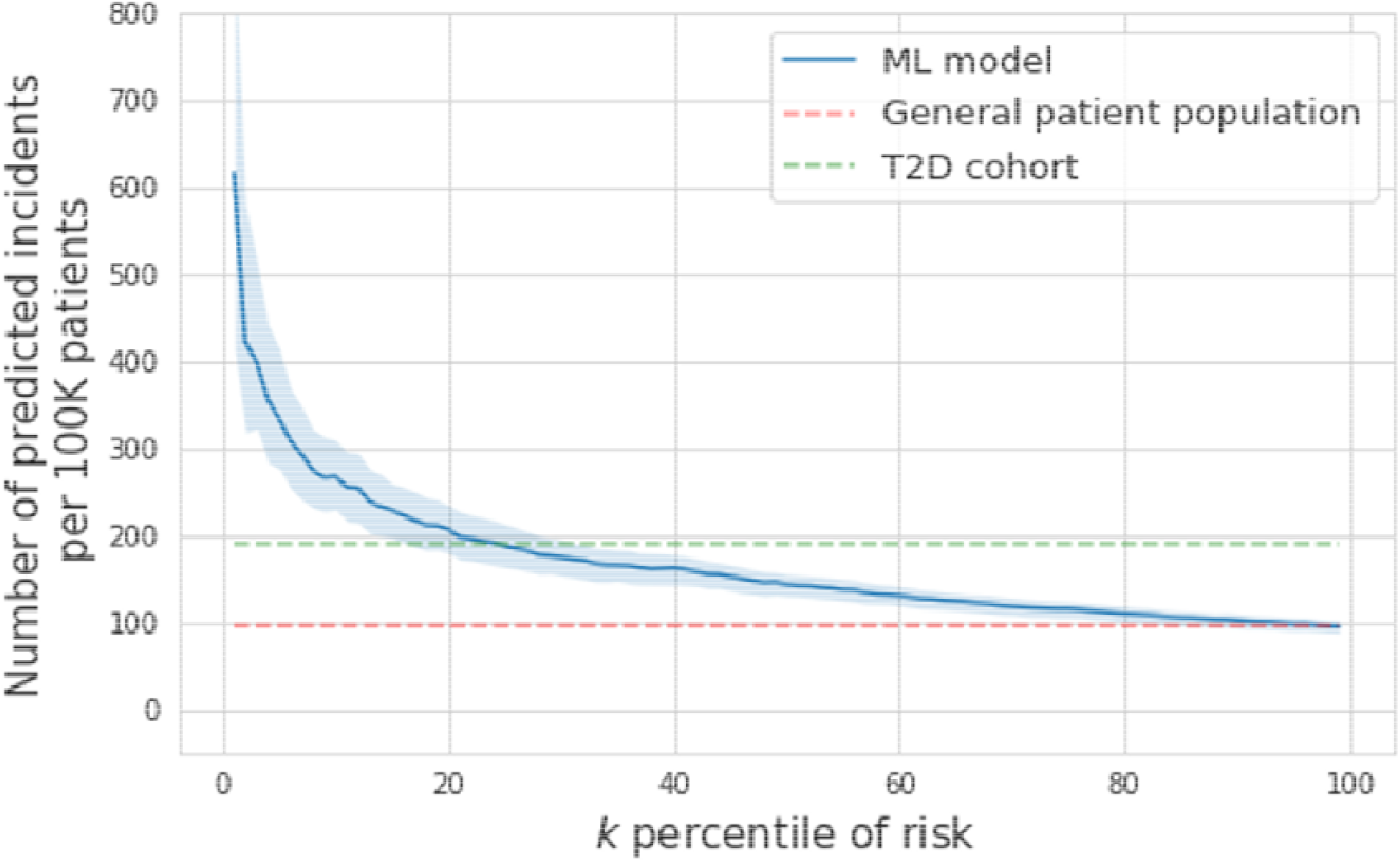
The positive predictive value (scaled to depict rate of identification in 100,000 patients), of risk of new-onset Pancreatic Cancer in 3-year follow-up, at different predicted risk levels.

### 3.4 Sensitivity Analysis

We conducted the sensitivity analysis with 10 trials that repeated training and validation with simulated datasets by randomly flipping outcomes. The AUC of simulated experiments was 0.718 [0.713, 0.722], which is not significantly different from the performance of the original dataset. The sensitivity analysis demonstrated that the model is robust under some noises in EHR.

## 4. Discussion

Although screening and early detection of pancreatic cancer hold a realistic promise to impact the survival of this disease, most patients diagnosed with cancer would not have been recommended to undergo screening. An important reason for this is the absence of strong risk factors that have precluded the identification of populations at risk [8,9]. Although high-risk populations have been identified based on familial or genetic risk, it is estimated that only a minority of patients diagnosed with pancreatic cancer have a currently identifiable predisposition [10,11]. Due to the absence of an effective screening tool or modality and the low prevalence (which is likely associated with many false positive results), pancreatic cancer screening in the general population is not recommended [12].

Despite the rapidly progressing and aggressive nature of pancreatic adenocarcinoma, there are multiple lines of evidence showing the time frame of progression from advanced precursors or early-stage cancer to advanced cancer around 1.5-3 years [13,14]. In addition, several biomarkers (i.e. CA19-9, glycemic indices, and others) show a significant change 12-36 months prior to the ultimate diagnosis of cancer [15–18]. Perhaps equally important is the observation that survival following the diagnosis and treatment of these late precursors or early cancers is greater than would be expected from lead time bias alone. We therefore hypothesized that recognizing patients at risk for pancreatic cancer approximately three years prior to a clinical diagnosis would be a biologically reasonable time frame that may allow diagnosis of high-grade precursors or early cancers and ultimately impact survival.

We leveraged the large electronic health record of our institution to identify a model that can select patients at risk for pancreatic cancer based on diagnostic variables, laboratory results, and demographic information. We evaluated the presence of these variables in a six-month time period 30 months prior to the cancer diagnosis and matched this cohort at a 1:4 ratio with a control group.

The strength of our approach is that it did not rely on any known association with conditions or diseases although we did limit the inclusion of laboratory tests to those that would have potential clinical relevance to the diagnosis of pancreatic diseases. In our variable selection, we ultimately identified 78 variables; 73 of which were diagnostic codes. The majority of these diagnostic codes had a known or plausible association with an individual risk factor for pancreatic cancer although only a few of them alone could potentially prompt screening for pancreatic cancer. One set of such variables was the diagnosis of certain pancreatic conditions such as pancreatic cysts and pancreatitis. We therefore excluded patients with a known history of pancreatic disease or a presumed evaluation that would’ve either identified pancreatic disease or confirmed the presence of a normal pancreas. However, ultimately this represented a small cohort and it had no major impact on model performance.

The area under the curve of our model was 0.742. Our precision-recall curve shows that using our model, the top 1% risk percentile is associated with a nearly 7 fold increased risk. As the accuracy of the screening instruments is limited (especially lacking the ability to detect precursor lesions), it is somewhat difficult to compare these numbers to other cancer screening. However, the rate of cancer diagnosis in colorectal cancer screening studies per patient in the general population ranges from 1:154-208 depending on the screening method [20,21] and is 1:250 in lung cancer screening by CT scan [22]. This would suggest that the highest risk individuals in the model would constitute a sufficiently enriched cohort to offer screening.

Our study has several limitations. The setting and health record we studied is that of an academic institution and its affiliates and our data is entirely limited to patients who continue to get their care in this system. An analysis on a much greater scale or external validation in a different demographic will be essential. Additionally, our variables are static and changes in metabolic parameters such as weight or glycemic control are not considered. Although this makes the model easier to implement, future approaches will likely need to consider the impact of change.

## 5. Conclusion

In conclusion, our study shows that it is feasible to identify a patient population at 10 fold higher risk for developing pancreatic cancer than the average population using static variables in a machine learning model. This approach offers an easily implementable approach toward finding an enriched population at sufficiently high baseline risk to warrant a discussion about screening. We believe that an EHR-based approach to identifying individuals coupled with a notification system _[SD1]_ has the chance to reduce disparities in screening and recognize novel high-risk populations. It is however critical to recognize that the current screening tests for pancreatic cancer have significant weakness, which will undoubtedly affect the benefit of finding a high-risk population. Yields of screening in current high-risk cohorts have been relatively modest and this may be due to selection based on only a few variables. Our future efforts will focus on measuring the impact of the implementation of this machine learning based model and this will hopefully occur parallel to continued improvements in the use of screening modalities.

## Data Availability

Due to protection of private patient information, the data used in the study will not be made publicly available on the basis of individual collaborations. If an institution-wide agreement between NYU Langone and the requesting institutions is generated, we will share the agreed data elements under data use agreement terms.

---

**Figure B1.**
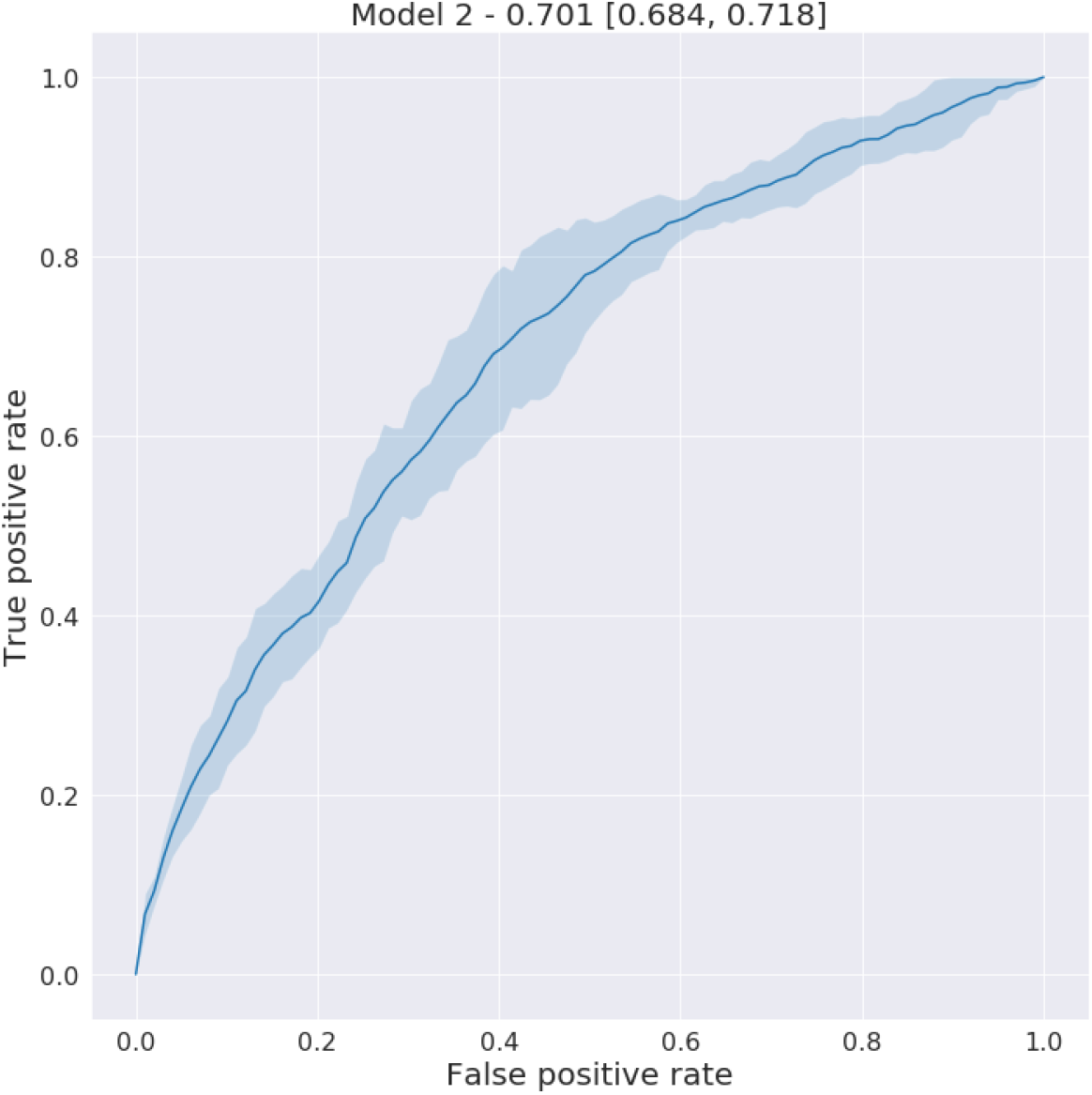
The Receiver Operating Characteristics Curve (AUROC 0.701 [0.684, 0.718]) of Model 2 on the held-out validation set to predict new-onset pancreatic cancer in a 3-year follow-up.

**Table A1.**
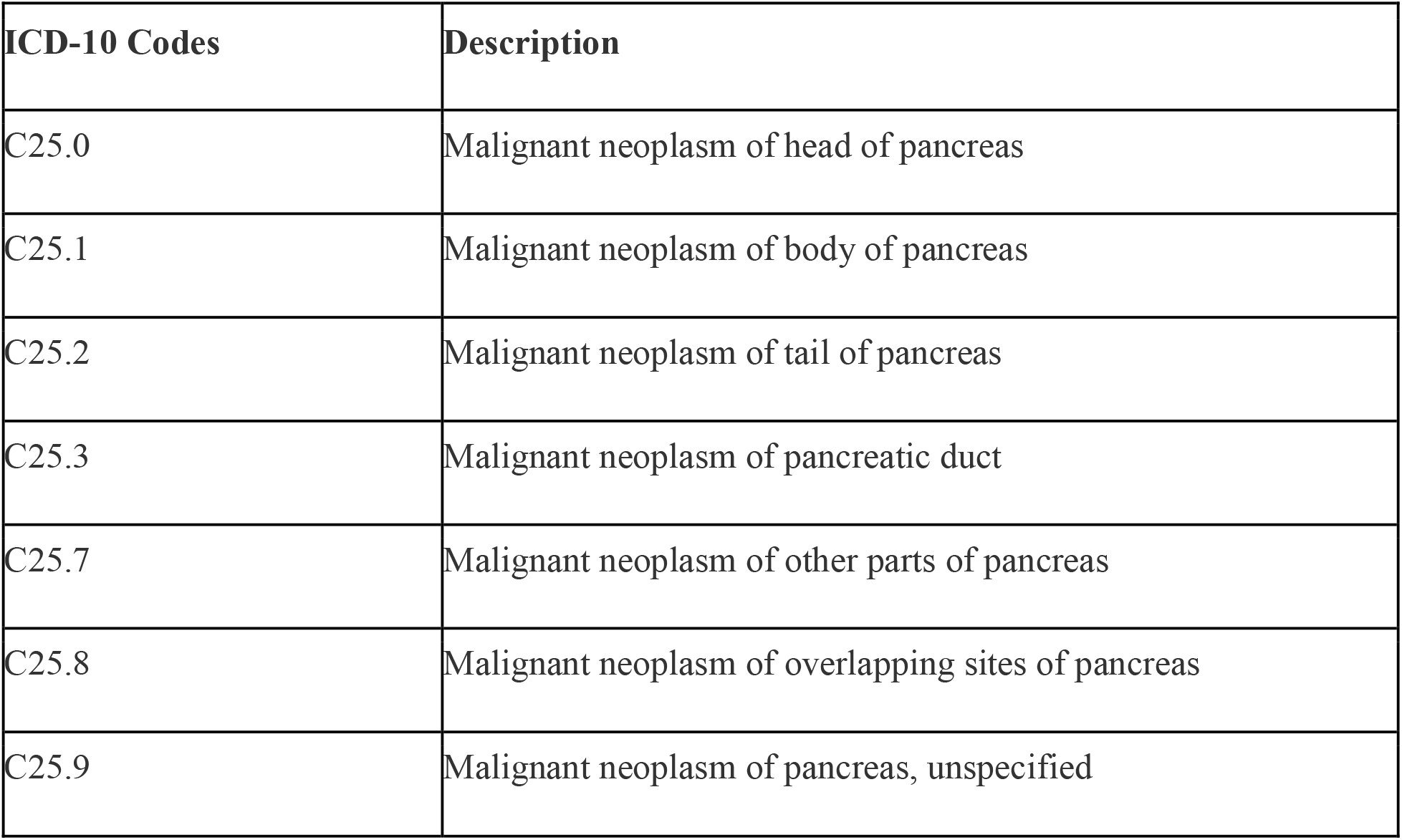
Definition of pancreatic cancers referring to ICD-10 codes.

| **ICD-10 Codes** | **Description** |
| --- | --- |
| C25.0 | Malignant neoplasm of head of pancreas |
| C25.1 | Malignant neoplasm of body of pancreas |
| C25.2 | Malignant neoplasm of tail of pancreas |
| C25.3 | Malignant neoplasm of pancreatic duct |
| C25.7 | Malignant neoplasm of other parts of pancreas |
| C25.8 | Malignant neoplasm of overlapping sites of pancreas |
| C25.9 | Malignant neoplasm of pancreas, unspecified |

**Table A2.** Normal ranges of lab values used in this study.

| **Lab** | **Threshold used for abnormal laboratory test** |
| --- | --- |
| A1C (Diabetes) | < 6.5% |
| CA19-9 | < 37 mg/dL |
| Lipase | < 234 U/L |
| Amylase | < 330 U/L |
| AST | < 40 |
| ALT | < 55 |
| *Albumin* | *< 3.4* |
| *Hemoglobin (HGB)* | *> 13 g/dL* |
| *Fasting glucose* | *< 126 mg/dL* |

**Table A3.** The full list of selected features by PheWAS. The unadjusted odds ratios on the case-control cohort and adjusted odds ratios of two models on the overall cohort are listed. *Note that the statistically significant features after bonferroni correction are marked (*** means corrected p-value < 0.001; ** means corrected p-value < 0.05; * means corrected p-value < 0.1)

| **Diagnosis Features** | **Control (%)** | **Case (%)** | **P-value** | **Unadjusted Odds Ratio**<br>**[95% Confidence Interval]** |
| --- | --- | --- | --- | --- |
| Disease of pancreas; unspecified (ICD K86.9) | 2(0.02%) | 11(0.53%) | 5.61E-09 | 22.11 [4.90 ; 99.83] |
| Malignant neoplasm of transverse colon (ICD C18.4) | 1(0.01%) | 5(0.24%) | 1.05E-04 | 20.05 [2.34 ; 171.68] |
| Other chronic pancreatitis ((ICD K86.1) | 2(0.02%) | 8(0.38%) | 2.08E-06 | 16.06 [3.41 ; 75.67] |
| Hemangioma of intra-abdominal structures (ICD D18.03) | 1(0.01%) | 4(0.19%) | 7.94E-04 | 16.03 [1.79 ; 143.49] |
| Malignant neoplasm of unspecified allopan tube (ICD C57.00) | 1(0.01%) | 4(0.19%) | 7.94E-04 | 16.03 [1.79 ; 143.49] |
| Cyst of pancreas (ICD K86.2) | 13(0.16%) | 46(2.20%) | 6.21E-29 | 14.45 [7.79 ; 26.80] |
| Peritoneal adhesions (postprocedural) postinfection) (ICD K66.0) | 1(0.01%) | 3(0.14%) | 5.95E-03 | 12.02 [1.25 ; 115.58] |
| Neoplasm of uncertain behavior of digestive organ; unspecified (ICD D37.9) | 1(0.01%) | 3(0.14%) | 5.95E-03 | 12.02 [1.25 ; 115.58] |
| Encounter for breast reconstruction following mastectomy (ICD Z42.1) | 1(0.01%) | 3(0.14%) | 5.95E-03 | 12.02 [1.25 ; 115.58] |
| (ICD I48.21) | 1(0.01%) | 3(0.14%) | 5.95E-03 | 12.02 [1.25 ; 115.58] |
| Body mass index (BMI) 39.0-39.9; adult (ICD Z68.39) | 1(0.01%) | 3(0.14%) | 5.95E-03 | 12.02 [1.25 ; 115.58] |
| Presence of urogenital implants (ICD Z96.0) | 1(0.01%) | 3(0.14%) | 5.95E-03 | 12.02 [1.25 ; 115.58] |
| Personal history of poliomyelitis (ICD Z86.12) | 1(0.01%) | 3(0.14%) | 5.95E-03 | 12.02 [1.25 ; 115.58] |
| Chronic superficial gastritis without bleeding (ICD K29.30) | 1(0.01%) | 3(0.14%) | 5.95E-03 | 12.02 [1.25 ; 115.58] |
| Pseudocyst of pancreas (ICD K86.3) | 6(0.07%) | 16(0.77%) | 6.04E-10 | 10.74 [4.20 ; 27.48] |
| Hypertrophy of breast (ICD10 N62) | 4(0.05%) | 9(0.43%) | 8.98E-06 | 9.03 [2.78 ; 29.37] |
| Aftercare following surgery for neoplasm (ICD Z48.3) | 4(0.05%) | 9(0.43%) | 8.98E-06 | 9.03 [2.78 ; 29.37] |
| Neoplasm of unspecified behavior of digestive system (ICD D49.0) | 8(0.10%) | 16(0.77%) | 1.05E-08 | 8.05 [3.44 ; 18.84] |
| Personal history of other malignant neoplasm of kidney (ICD Z85.528) | 3(0.04%) | 6(0.29%) | 4.63E-04 | 8.02 [2.00 ; 32.10] |
| Acute frontal sinusitis; unspecified (ICD J01.10) | 2(0.02%) | 4(0.19%) | 4.26E-03 | 8.01 [1.47 ; 43.78] |
| Body mass index (BMI) 31.0-31.9; adult (ICD Z68.31) | 2(0.02%) | 4(0.19%) | 4.26E-03 | 8.01 [1.47 ; 43.78] |
| Personal history of other malignant neoplasm of rectum; rectosigmoid junction; and anus (ICD Z85.048) | 4(0.05%) | 7(0.33%) | 2.95E-04 | 7.02 [2.05 ; 24.00] |
| Incomplete uterovaginal prolapse (ICD N81.2) | 3(0.04%) | 5(0.24%) | 2.64E-03 | 6.68 [1.60 ; 27.98] |
| Hypertrophy of nasal turbinates (ICD J34.3) | 3(0.04%) | 5(0.24%) | 2.64E-03 | 6.68 [1.60 ; 27.98] |
| Malignant neoplasm of ascending colon (ICD C18.2) | 3(0.04%) | 5(0.24%) | 2.64E-03 | 6.68 [1.60 ; 27.98] |
| Other abnormal tumor markers (ICD R97.8) | 5(0.06%) | 8(0.38%) | 1.79E-04 | 6.42 [2.10 ; 19.65] |
| (ICD C20) | 4(0.05%) | 6(0.29%) | 1.56E-03 | 6.01 [1.70 ; 21.33] |
| Body mass index (BMI) 37.0-37.9; adult (ICD Z68.37) | 4(0.05%) | 6(0.29%) | 1.56E-03 | 6.01 [1.70 ; 21.33] |
| Other specified diseases of pancreas (ICD K86.89) | 11(0.13%) | 15(0.72%) | 1.50E-06 | 5.49 [2.52 ; 11.96] |
| Genetic susceptibility to other malignant neoplasm (ICD Z15.09) | 10(0.12%) | 13(0.62%) | 1.17E-05 | 5.23 [2.29 ; 11.94] |
| Benign lipomatous neoplasm of other sites (ICD D17.79) | 4(0.05%) | 5(0.24%) | 7.63E-03 | 5.01 [1.34 ; 18.67] |
| Encounter for surgical aftercare following surgery on the digestive system (ICD Z48.815) | 4(0.05%) | 5(0.24%) | 7.63E-03 | 5.01 [1.34 ; 18.67] |
| Genetic susceptibility to malignant neoplasm of breast (ICD Z15.01) | 9(0.11%) | 11(0.53%) | 8.97E-05 | 4.91 [2.03 ; 11.86] |
| Malignant neoplasm of unspecified ovary (ICD C56.9) | 6(0.07%) | 7(0.33%) | 2.27E-03 | 4.68 [1.57 ; 13.94] |
| Genetic susceptibility to malignant neoplasm of ovary (ICD Z15.02) | 8(0.10%) | 9(0.43%) | 6.78E-04 | 4.52 [1.74 ; 11.72] |
| Benign neoplasm of ascending colon (ICD D12.2) | 9(0.11%) | 9(0.43%) | 1.45E-03 | 4.01 [1.59 ; 10.12] |
| Acute pancreatitis without necrosis or infection; unspecified (ICD K85.90) | 7(0.08%) | 7(0.33%) | 4.98E-03 | 4.01 [1.41 ; 11.45] |
| Left upper quadrant pain (ICD R10.12) | 7(0.08%) | 7(0.33%) | 4.98E-03 | 4.01 [1.41 ; 11.45] |
| Other specified personal risk factors; not<br>elsewhere classified (ICD Z91.89) | 27(0.32%) | 26(1.24%) | 1.15E-07 | 3.89 [2.26 ; 6.68] |
| Dependence on renal dialysis (ICD<br>Z99.2) | 8(0.10%) | 7(0.33%) | 9.77E-03 | 3.51 [1.27 ; 9.69] |
| Personal history of other endocrine;<br>nutritional and metabolic disease (ICD<br>Z86.39) | 8(0.10%) | 7(0.33%) | 9.77E-03 | 3.51 [1.27 ; 9.69] |
| Thoracic aortic aneurysm; without<br>rupture (ICD I71.2) | 13(0.16%) | 11(0.53%) | 1.54E-03 | 3.40 [1.52 ; 7.59] |
| Family history of malignant neoplasm;<br>unspecified (ICD Z80.9) | 18(0.22%) | 15(0.72%) | 2.51E-04 | 3.35 [1.69 ; 6.66] |
| Primary osteoarthritis; unspecified hand<br>(ICD M19.049) | 10(0.12%) | 8(0.38%) | 9.46E-03 | 3.21 [1.26 ; 8.14] |
| Other specified counseling (ICD Z71.89) | 23(0.27%) | 18(0.86%) | 1.26E-04 | 3.15 [1.70 ; 5.85] |
| Malignant neoplasm of unspecified<br>kidney; except renal pelvis (ICD C64.9) | 14(0.17%) | 10(0.48%) | 7.89E-03 | 2.87 [1.27 ; 6.46] |
| (ICD R69) | 17(0.20%) | 12(0.57%) | 3.95E-03 | 2.83 [1.35 ; 5.94] |
| Long term (current) use of insulin (ICD<br>Z79.4) | 43(0.51%) | 29(1.39%) | 1.59E-05 | 2.72 [1.70 ; 4.37] |
| Other specified diseases of liver (ICD<br>K76.89) | 30(0.36%) | 20(0.96%) | 3.94E-04 | 2.68 [1.52 ; 4.73] |
| Cerebral infarction due to unspecified<br>occlusion or stenosis of unspecified<br>cerebral artery (ICD I63.50) | 24(0.29%) | 15(0.72%) | 3.88E-03 | 2.51 [1.31 ; 4.79] |
| Nausea (ICD R11.0) | 34(0.41%) | 21(1.00%) | 7.25E-04 | 2.49 [1.44 ; 4.29] |
| Family history of malignant neoplasm of other organs or systems (ICD Z80.8) | 21(0.25%) | 13(0.62%) | 7.75E-03 | 2.49 [1.24 ; 4.97] |
| Disease of stomach and duodenum; unspecified (ICD K31.9) | 28(0.33%) | 17(0.81%) | 2.81E-03 | 2.44 [1.33 ; 4.47] |
| Monoclonal gammopathy (ICD D47.2) | 24(0.29%) | 14(0.67%) | 9.31E-03 | 2.34 [1.21 ; 4.54] |
| Other obesity due to excess calories (ICD E66.09) | 24(0.29%) | 14(0.67%) | 9.31E-03 | 2.34 [1.21 ; 4.54] |
| Other diseases of stomach and duodenum (ICD K31.89) | 55(0.66%) | 31(1.48%) | 1.87E-04 | 2.27 [1.46 ; 3.54] |
| Overweight (ICD E66.3) | 43(0.51%) | 24(1.15%) | 1.16E-03 | 2.25 [1.36 ; 3.71] |
| Epigastric pain (ICD R10.13) | 72(0.86%) | 38(1.82%) | 1.26E-04 | 2.13 [1.43 ; 3.17] |
| Family history of malignant neoplasm of digestive organs (ICD Z80.0) | 63(0.75%) | 33(1.58%) | 4.04E-04 | 2.11 [1.38 ; 3.23] |
| Type 2 diabetes mellitus with hyperglycemia (ICD E11.65) | 56(0.67%) | 29(1.39%) | 1.09E-03 | 2.09 [1.33 ; 3.28] |
| Unspecified abdominal pain (ICD R10.9) | 144(1.72%) | 63(3.01%) | 1.50E-04 | 1.77 [1.31 ; 2.39] |
| Malignant neoplasm of unspecified site of unspecified female breast (ICD C50.919) | 101(1.21%) | 43(2.06%) | 2.89E-03 | 1.72 [1.20 ; 2.46] |
| Constipation; unspecified (ICD K59.00) | 111(1.33%) | 45(2.15%) | 5.39E-03 | 1.64 [1.15 ; 2.32] |
| Mixed hyperlipidemia (ICD E78.2) | 228(2.73%) | 89(4.26%) | 2.62E-04 | 1.59 [1.24 ; 2.04] |
| Type 2 diabetes mellitus without complications (ICD E11.9) | 459(5.49%) | 164(7.84%) | 4.71E-05 | 1.47 [1.22 ; 1.76] |

